# Exploring Causal relationship between risk factors and vulnerability to COVID-19 Cases of Italy, Spain, France, Greece, Portugal, Morocco and South Africa

**DOI:** 10.1101/2020.06.24.20139121

**Authors:** Abdelghani Youmni, Mbarek Cheikh

## Abstract

Even though the infection rate of COVID-19 is very high as of today 31 May: 5,819,962 confirmed cases worldwide, the death rate is only about 6.23%, 362,786 deaths as for the same date. Furthermore, the rate of total infected cases is extremely different from one country to another as well as for the rate of mortality. Therefore, there may be some factors that possibly amplify the rate of infection from one country to another as well as for the rate of mortality due to COVID-19. In the literature, we have found multiple identified risk factors responsible for vulnerability to COVID19, we have chosen pertinent key risk factors for our study: Median-age, age>65 years old, weight, population density, diabetics, International arrivals, median temperature between March and May.

**Objective:** We aim to find correlation between the identified risk factors and vulnerability to COVID-19 in seven different countries from Europe and Africa: Most affected countries Italy, Spain, France, moderately affected: Portugal, and less affected countries: Greece, Morocco and South Africa.

**Data sources:** WHO, Worldometers, Ourworldindata

**Population:** all reported COVID-19 total in-hospital infected and death cases in Italy, Spain, France, Portugal, Greece, Morocco, and South Africa.

**Time period:** 15^th^ March 2020 to 15^th^ May 2020

**Methods:** We used Multiple linear regression in our approach to modeling the relationship between the dependent variable **(DV)**: vulnerability to COVID19 (which we presented by number of totals in-hospital infected cases per million for each country) and the independent variables **(IV)** scores: median age, aged 65+, population density, international arrivals, BMI, diabetes prevalence, and temperature. We used SPSS software to generate multiple linear regression; Pearson correlation factor: r, ANOVA table, Coefficients table, as well as bar charts and scatter plots.

The multiple linear regression equation of our study model is:

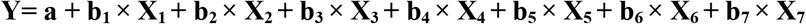

Where:

Y: Predicted score on total cases per million or Vulnerability to COVID19; a: intercept; b1: regression coefficient or weight for median age; X1: median age; b2: regression coefficient or weight for population density; X2: population density; b3: regression coefficient or weight for international arrivals; X3: international arrivals in millions; b4: regression coefficient or weight average temperature; X4: average temperature in Celsius; b5: regression coefficient or weight for BMI; X5: BMI Body Mass Index in Kg/m; b6: regression coefficient or weight for diabetes prevalence percentage; X6: diabetes prevalence percentage; B7: regression coefficient or weight for aged 65+; X7: aged 65+ percentage

**Results:** Till 15th May 2020 There were in Spain: 229540 total infected cases and 27321 total death attributed to COVID-19, France: 141356 total cases and 27425 total deaths, Italy: 223096 total cases and 31368 total deaths, Portugal: 28319 total cases and 1184 total deaths, Greece: 2770 total cases and 156 total deaths, Morocco: 6607 total cases and 190 total deaths, and South Africa: 12739 total cases and 238 total deaths. In summary after full adjustment, total death cases were strongly associated with total infected cases for the population of all the seven chosen countries combined: correlation factor r= 0.921 with a P-value= .000: P<0.05 (Sig.(2-tailed). Population density was significantly correlated with total infected cases for all the seven countries combined: r= 0.478 with a P-value= .000: P<0.05. the median age was moderately associated with infected cases: r= 0.563 with P<0.05. However, diabetes prevalence was less associated with infected cases: r= 0.146 with P<0.05. International exposure was significantly associated with infected cases: r= 0.609 with P<0.05. Median temperature was negatively correlated with total infected cases in the seven countries combined.

**Conclusions:** We have quantified a range of risk factors for infection and death from COVID-19, in seven highly, moderate and less affected countries, in a large correlation-regression study. People from countries with high international exposure, high population density, are at markedly risk of getting infected such as France, Spain, and Italy. Moreover, people from countries with high median age are at increased risk of in-hospital death from COVID19. Furthermore, by conducting multiple linear regression analysis we have found that the overall regression model is significant (as shown in the tables). 66.2% (r^2^=.662) of the vulnerability is explained by the model which includes all predictor variables or risk factors.

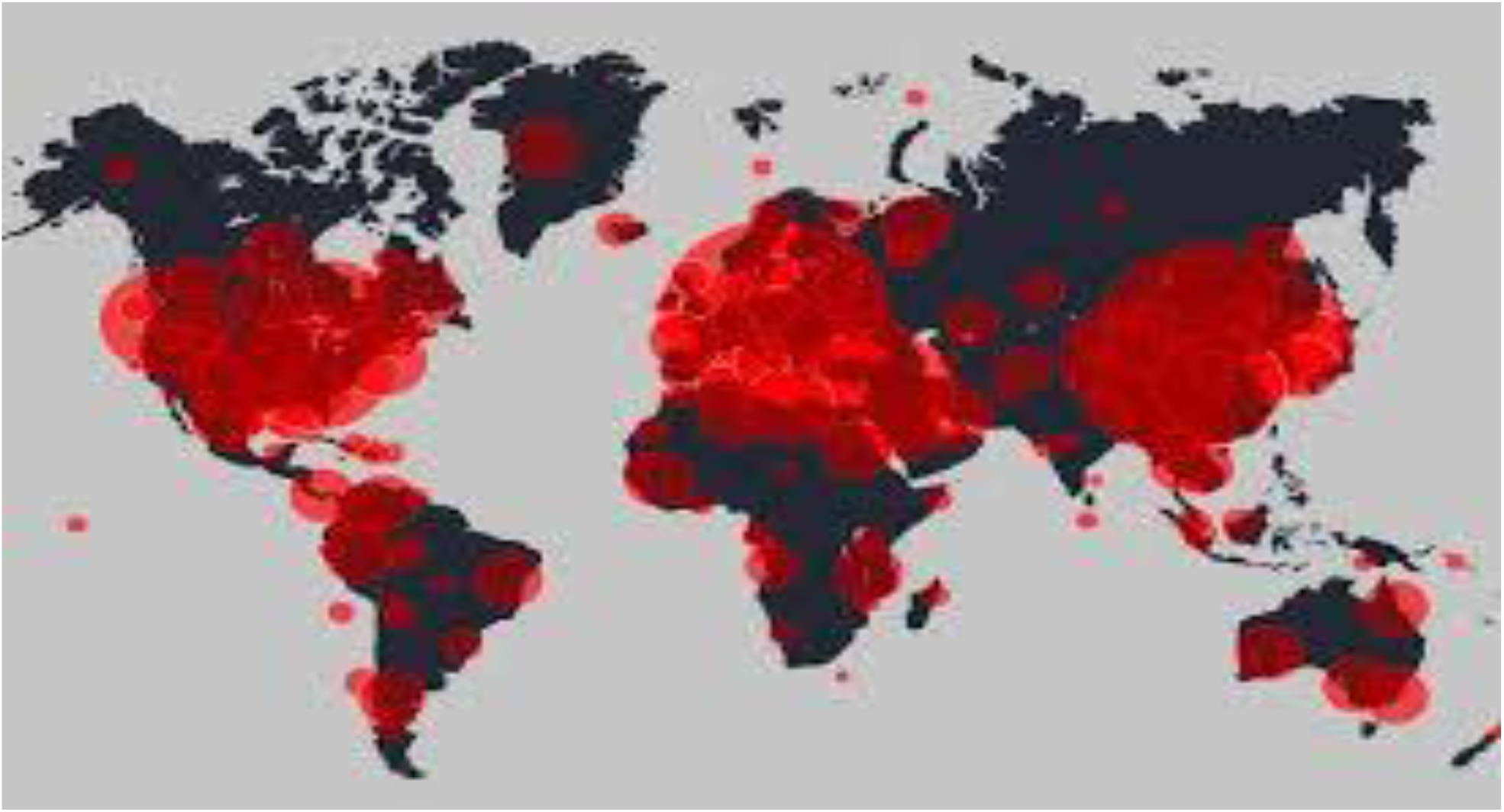

## INTRODUCTION

On 31 December 2019, Chinese authorities alerted the World Health Organization (WHO) to an outbreak of unknown etiology pneumonia in the city of Wuhan, Hubei province, China. As of January 22, 2020, more than 600 cases have been confirmed, including 444 from Hubei province^**1**^. COVID-19 has been affirmed to be transmissible from human-to-human^**2**^, which raised high attention not only in China but internationally. On March 11th 2020, the World Health Organization characterized COVID-19 as a pandemic after 118,000 cases and 4,291 deaths were reported in 114 countries. As of 15 May, 4,347,936 cases are over 4 million globally, with more than 307,000 deaths attributed to the virus. In Spain, cases have reached 229,540, with 27, 321deaths in hospital. France: 141356 total cases and 27425 total deaths, Italy: 223096 total cases and 31368 total deaths, Portugal: 28319 total cases and 1184 total deaths, Greece: 2770 total cases and 156 total deaths, Morocco: 6607 total cases and 190 total deaths, and South Africa: 12739 total cases and 238 total deaths^**3**^.

To test the correlation between vulnerability due to COVID-19 and the risk factors we have chosen some relevant risk factors based on several Chinese, European and American studies and publications. To do this, We used Multiple linear regression in our approach to modeling the relationship between the dependent variable (DV): vulnerability to COVID19 (which we presented by number of totals in-hospital infected cases per million for each country) and the independent variables, probable risk factors approved by other studies, which are: median age, aged 65+, population density, international arrivals, BMI, diabetes prevalence, and temperature.

Age, population density, temperature, and International Exposure are well-established risk factors, with over r = .50 correlation. Age, as consistent with global patterns, is associated with increased risk of death. However, Weight and diabetes are not statistically significantly correlated with affected cases. These factors often correlate with age, but correction for age was not possible in the available data.

## METHODS

### Study design

We conducted multiple linear regression in our approach to modeling the relationship between the dependent variable: vulnerability to COVID19 (which we presented by number of totals in-hospital infected cases per million for each country) and the independent variables scores: median age, aged 65+, population density, international arrivals, BMI, diabetes prevalence, and temperature. We used SPSS software to generate multiple linear regression, Pearson correlation factor: r, ANOVA table, Coefficients table, as well as graphs like scatter plots.

The multiple linear regression equation of our study model is:

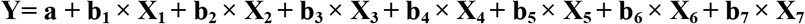

Where:

A correlation-regression study using World Health Organization (WHO) health record data linked to in-hospital COVID-19 infected and death data (see Data Source). The study began on 15 March 2020 chosen as a date where most countries started sanitary measures such as lockdown several weeks after the first reported COVID-19 deaths in Morocco for instance; and ended on 15th May 2020. All the statistical analyses were two-sided at a 5% level of significance. All analyses were conducted using SPSS software (version 26). The correlation-multiple linear regression study explores causal relationship between risk factors and infected cases among the general infected in-hospital population in the seven countries.

### Data Source

We retrieved our data from COVID-19 infected hospitalized cases data available on WHO official website, Ourworldindata site, and worldometers records from March 15th to May 15th 2020, managed by the SPSS software, version 26.

### Outcomes

The outcome was in-hospital confirmed infected COVID-19 cases as well as death among people with confirmed COVID-19,

### Covariates

Potential risk factors included: Age, body mass index (BMI; kg/m2, Diabetes prevalence, International arrivals, Population Density and median temperature in March and May. Where categorized, age groups were: Obesity was grouped using categories derived from the World Health Organization classification of BMI: no evidence of obesity <30 kg/m obese I 30-34.9; obese II 35-39.9; obese III 40+.

## RESULTS

Numerous studies have shown that diabetes disease is an aggravating factor in COVID-19 infection. They Firstly reported the clinical features of 41 confirmed patients, and indicated that (32%) of them had underlying diseases^4^, including cardiovascular disease, diabetes, hypertension, and chronic obstructive pulmonary disease. In our study including five European countries and two African, the prevalence of diabetes is almost similar and the number of people with diabetes is on average 6.4%. We have noticed in several publications^**5**^ that elderly persons, men, and those with pre-existing hypertension and/or diabetes were highly prevalent in this case series and the pattern was similar to data reported from China. Among the patients who died, those with diabetes were more likely to have received invasive mechanical ventilation or care in the ICU compared with those who did not have diabetes^6^. In addition to these considerations, it should be noted that the seven countries on our panel have a temperate climate and a steady turnover of seasons, the model shows an interdependence between the vulnerability of the affected cases, the spread of the virus and the temperature variations associated with changing seasons.

**Graph.1.**
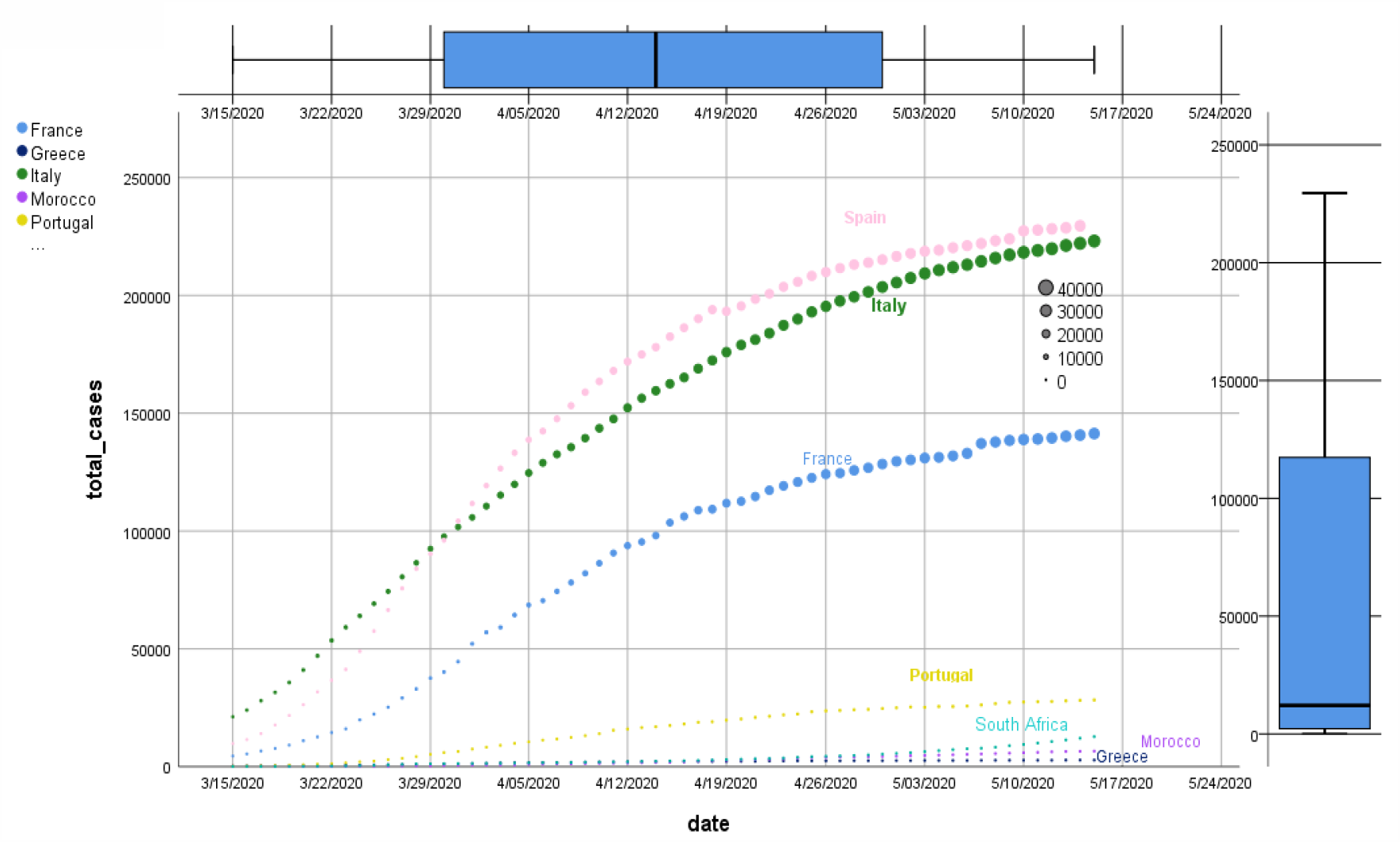

Graph.1 shows the distributions and development of the total infected in-hospital cases, and total death in-hospital cases for each country from 15 of March 2020 to 15 of May 2020. From the graph and until the 15 of March we can see that Spain and Italy have scored both more than 200000 infected Covid-19 cases, and France (about to reach 150000 infected cases) are the most affected countries in our study panel with a death rate more than 30000 cases for each country. However, Portugal was moderately affected with approximately 25000 infected cases; South Africa, Morocco, and Greece are the least affected countries.

**Graph.2.**
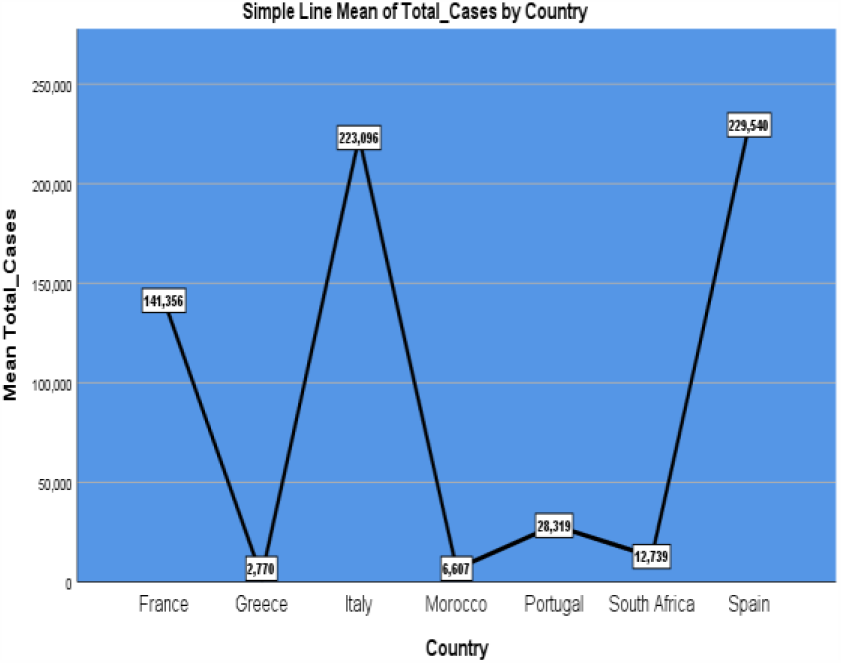

**Graph.3.**
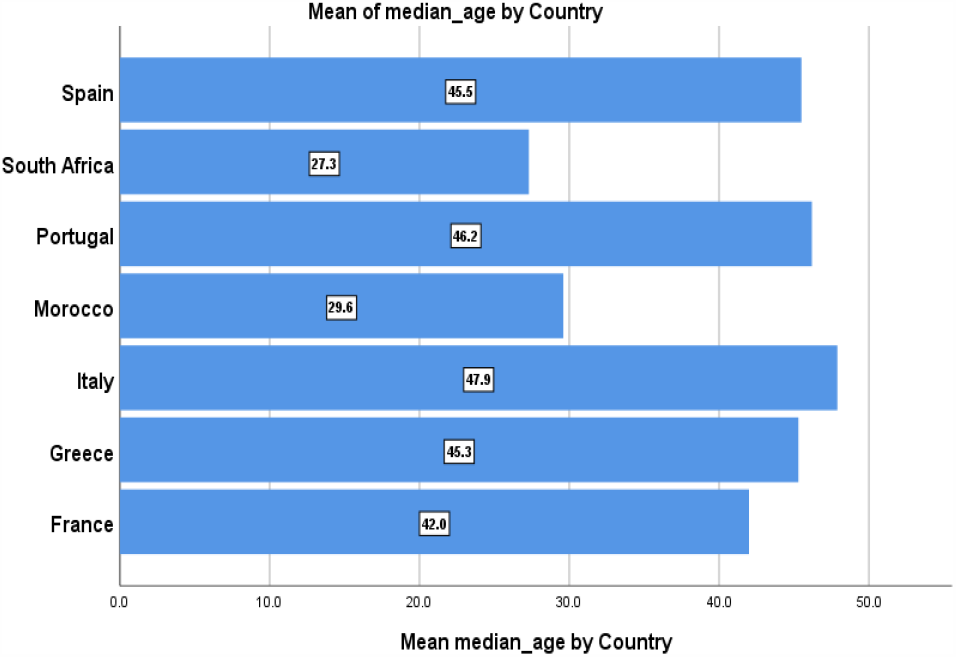

Age is a well-established risk factor correlated with death due to with over 90% of UK COVID-19 in-hospital deaths to date being in people aged over 60 years^7^, consistent with global patterns. In **graph**.**2** we can see the distribution of total cases per country till 15^th^ of May. Greece with only 2770 total infected cases contrary to Spain with 229540 total infected cases till the same date. Obviously, age is a well-established risk factor in many other studies. But, considering age alone can’t explain the big difference in total infected cases between Spain and Greece for instance (the former is 20 times affected than the latter) since both countries they have the same median age (45 years old) **graph**.**3**. Therefore, there may be other risk factors that may correlate with age.

**Graph.4.**
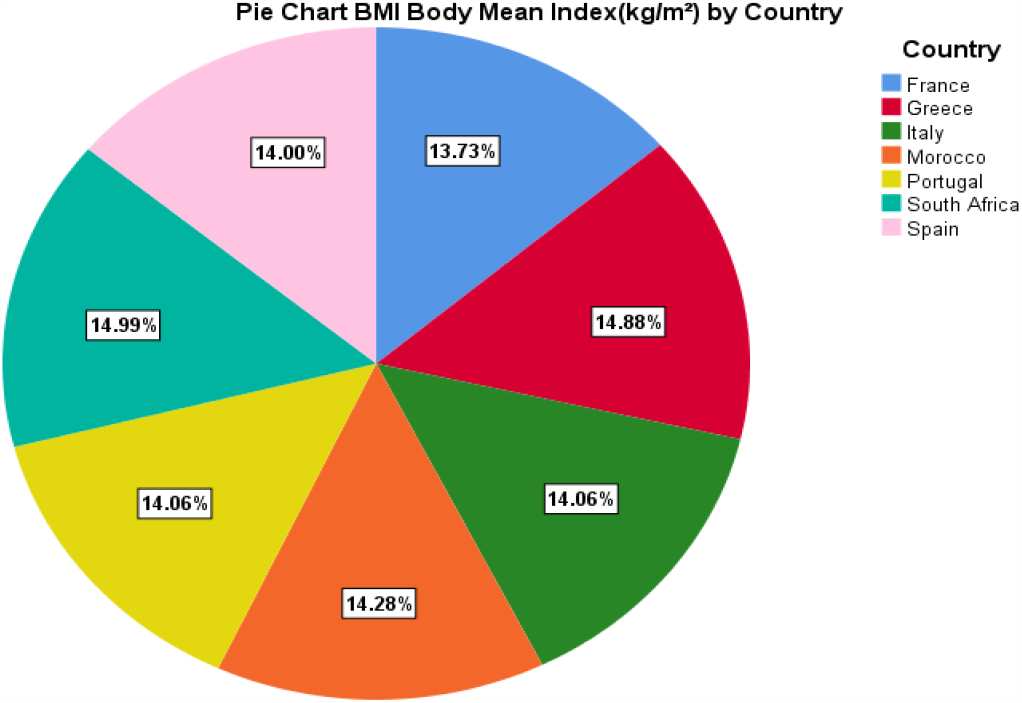

The pie Chart in **graph**.**4** we can notice that almost all the seven countries included in this study score approximately the same BMI (14 kg/m) which belongs to a category derived from the World Health Organization classification of BMI of no evidence of obesity <30 kg/m.

**Graph.5.**
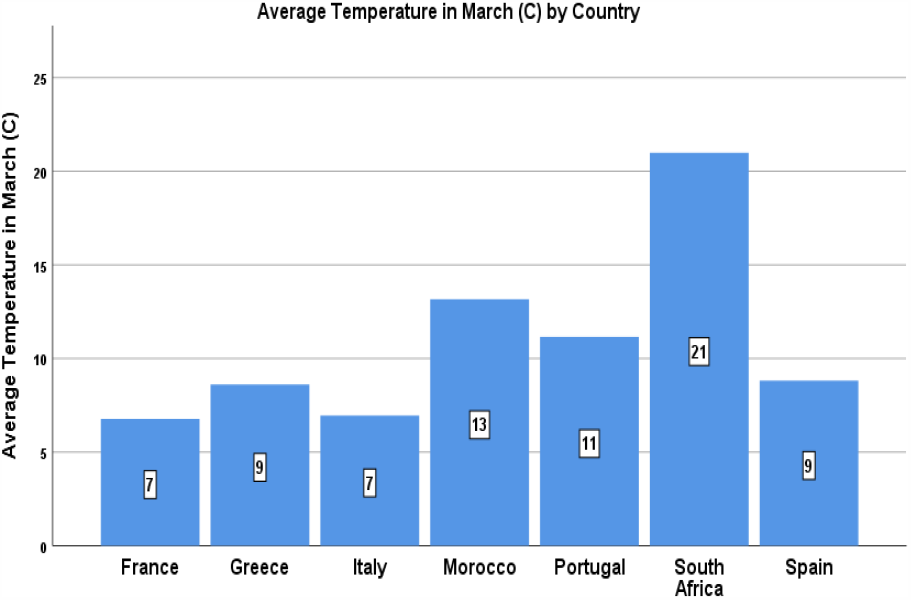

**Graph.6.**
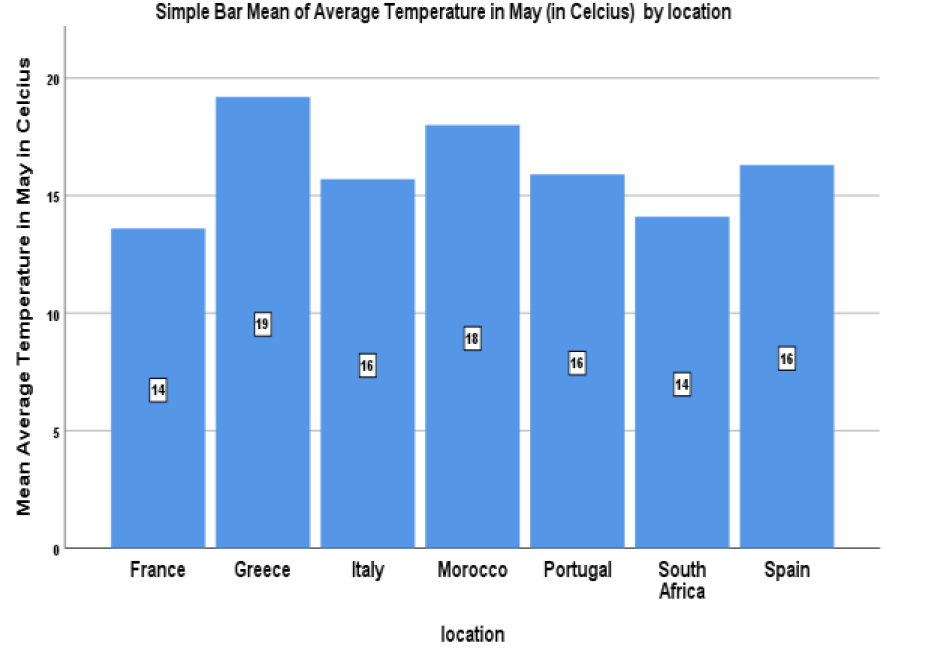

In **graph**.**5** we see that the average temperature in March in Morocco (13 C is uite the double in rance and taly C) and the average temperature scored in South Africa is the triple (21 C). However, In May (see **graph**.**6)** the average tem erature in outh Africa is only the same as in rance. orocco and reece almost scored the same tem erature and while taly Spain, and Portugal scored the same average tem erature in ay. rom the shar decrease of temperature between March and May in South Africa, we can anticipate an increase of vulnerability to COVID-19. On the other hand, we can anticipate a significant decrease in total in-hospital infected cases in the countries where the temperature has marked a sharp increase between March and May such as France, Spain, Italy.

**Graph.7.**
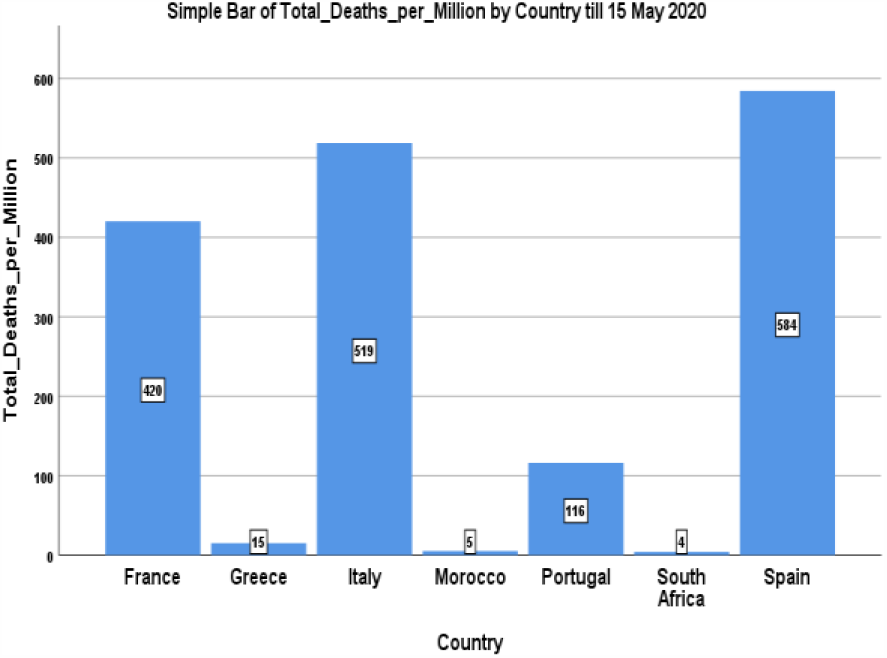

**Graph.8.**
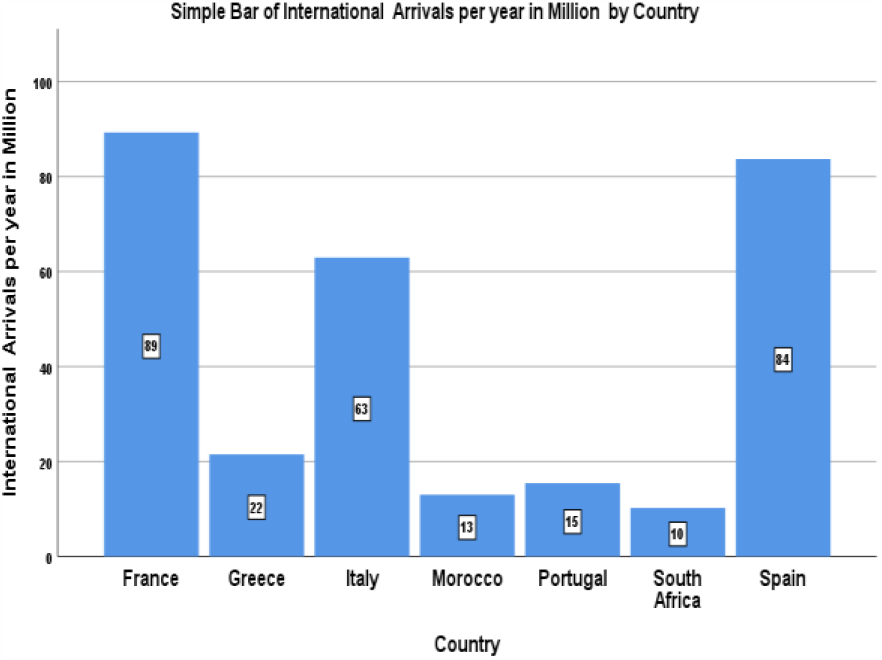

**Figure**.**7** shows the total deaths per million for each country till 15^th^ of May 2020 where Morocco and South Africa are scoring the least (5 and 4 deaths per million). Spain, Italy, and France in correlation with total cases rate are scoring highest death rate (584, 519, and 420 death per million). Here we can anticipate the correlation that may exist between age, total cases, and total deaths. **Graph**.**8** a bar chart of the international arrivals per year in millions for each country, also and based on the data we can anticipate a possible association between total number of infected cases and international openness. For instance, most infected countries Spain, Italy and France are the ones that scored the most in terms of international arrivals with 84, 63, and 89 million per year. Alternatively, the countries with least infected cases scored the least international arrivals: South Africa with 10 million and Morocco with 13 million arrivals per year.

**Graph.9.**
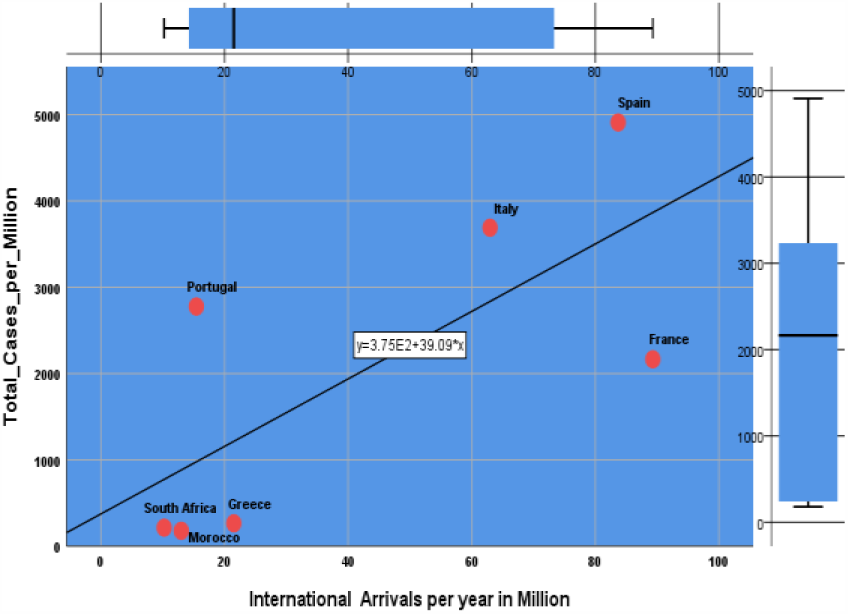

**Graph.10.**
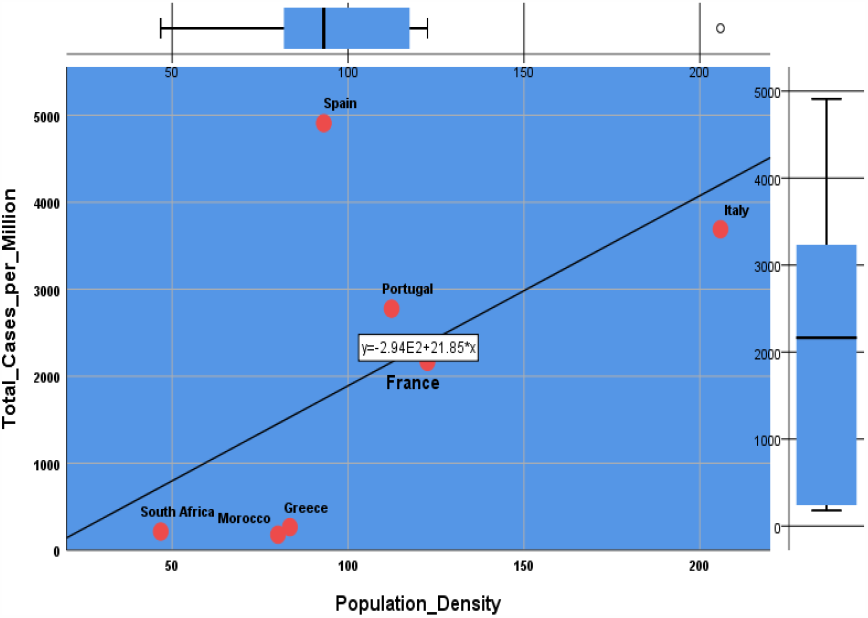

Population density **(graph**.**9):** The sign associated with the regression coefficient obtained is positive. This impact seems to be explained by the promiscuity, for high density clusters, such as crowds at mass gatherings, the random movement and contact of individuals initially increases the frequency of contacts. However, the contact rate decreases at extremely low density when it becomes difficult for people to move and make contact with others^8^. Scatter plot in **Graph**.**10** shows the positive correlation, in different degrees, between popualtion density and total affected cases for each country: Italy with more than 200 inhabitant per km^2^ is one of the most affected countries. South Africa with less than 50 inhabitants per Km^2^ is also one of the least affected countries. However, Spain with less 100 per Km^2^ nation wide is the most affected, but there are some cities in Spain with high popualtion density such as Madrid and Barcelone.

In this multitude of interdependencies between vulnerability and certain risk factors, the impact of travelers from abroad is also positive. The number of infected cases is directly correlated with number of air traffic routes and passenger volume. Therefore, continuous flight services from these secondary epicenters have presumably played a major role in COVID-19 spread^9^. The expected force of infection (AEF) powerfully explains pandemic risk, showing correlation of 0.90 to the transmission level needed to give a disease pandemic competence, and correlation of 0.85 to the delay until an outbreak becomes a pandemic^10^. **Graph**.**9** is a scatter plot that shows the positive correlation between international arrivals and total cases per million for each country with regression line and equation for all the countries coumbined. For instance, for each 1 million arrivals there may be 44 infected cases per million residents.

**Graph.11.**
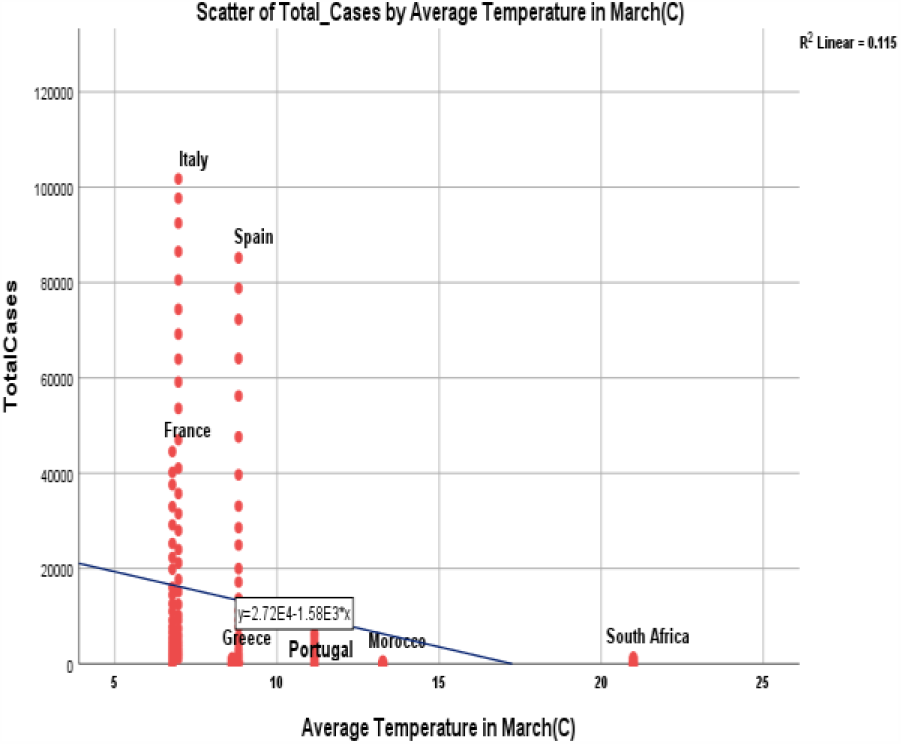

**Graph.12.**
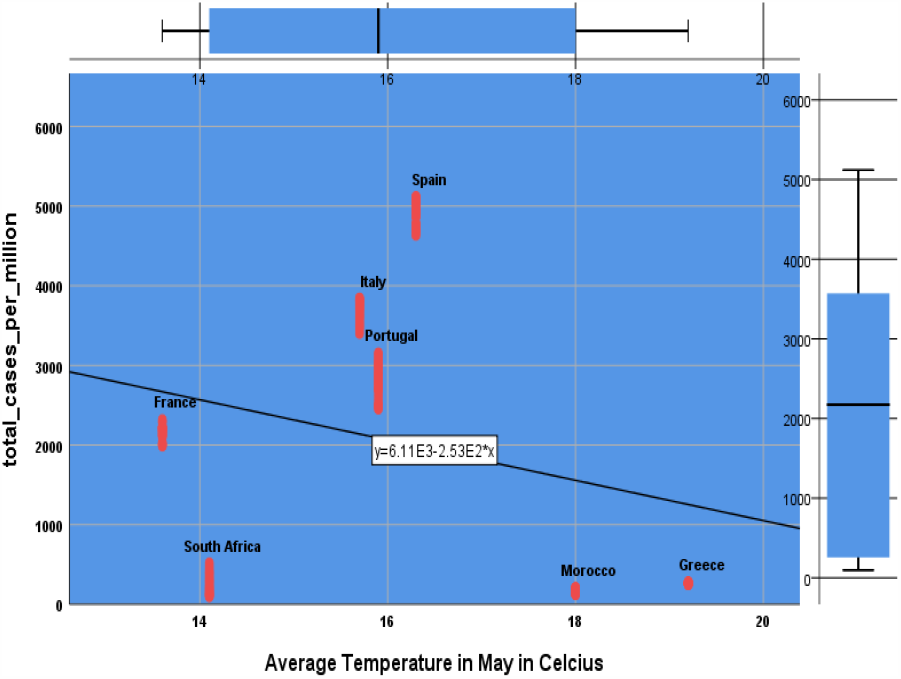

In related previous studies, the outbreak of severe acute respiratory syndrome (SARS) in Guangdong in 2003 has loosened gradually as weather temperature rises, and was basically ended till July^**11**^. Furthermore, it has been documented that the temperature and its variations might have affected the SARS outbreak^**12**^. A study in Korea found that the risk of influenza incidence was significantly increased with low daily temperature^**13**^. Moreover, temperature^**14**^ has been linked to the death from respiratory diseases. **Graph**.**11** confirms the negative correlation (a= −1.58) between temperature and total cases already proved in other studies.

The average monthly temperature in March is: 8.81°C in Spain, 6.77°C in France, 8.61°C in Greece, 6.96°C in Italy, 13.26°C in Morocco, 11.15°C in Portugal, and 20.99°C in South Africa. In accordance with what we anticipated, and due to the sharp decrease of temperature between March and May in South Africa, it scored more total infected in-hospital cases per million, see **graph**.**12** in May compared with Morocco where temperature is still increasing since March.

**Graph.13.**
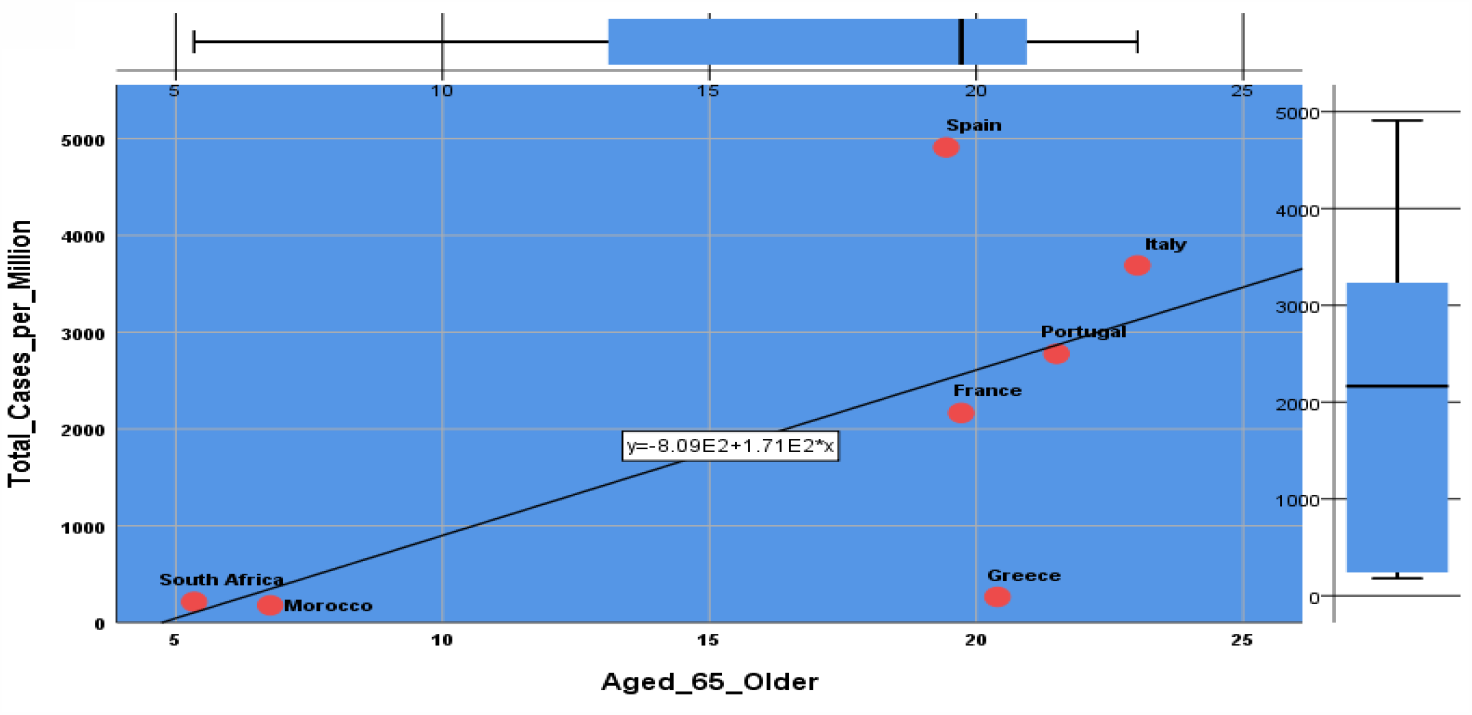

Aged above 65 years seems to be a determining factor in the vulnerability, hospitalization and death of individuals. The age pyramid in the seven countries on our panel shows real differences. We also evaluated the relation between vulnerability and Median age, we concluded that this parameter is significantly affecting people affected by the COVID-19; the correlation is positive and strong. The Center for Infectious Disease Research and Policy published in a study that on average, patients were middle-aged (median, 56 years), patients who died were, on average, 69 years old, versus 52 years in survivors^15^. When we contrast the figures of the seven countries, we note that the more than 65 years have on average 21.5% of the population in the five European countries of our panel and only 7% of the Moroccan population and 5% of South Africans.

**In graph**.**13** we see the positive strong correlation between the percentage of the people aged more than 65 years old and total in-hospital infected cases for almost all the countries except Greece which scored less than 200 per million even if it has more than 20% of its population are aged more than 65 years old.

### Multiple linear regression outputs using SPSS

Dependent variable: **Vulnerability to COVID-19 (score expressed by total infected in-hospital cases per million) in France, Spain, Italy, Portugal, Greece, Morocco, and South Africa** Independent variables:

‐ **Median Age score**,
‐ **Population density (number of inhabitants per Km**^**2**^),
‐ **International Arrivals in millions**,
‐ **Average temperature in Celsius**,
‐ **BMI Body Mass Index in Kg/m**
‐ **Diabetes prevalence percentage**
‐ **Aged 65+ percentage**

### Model Summary table

The overall regression model is significant (as shown below). 66.2% (r^2^=.662) of the variation is explained by the model which includes all predictor variables.

### Model Summary

| Model | R | R Square | Adjusted R Square | Std. Error of the Estimate | Change Statistics |  |  |  | Sig. F Change |
| --- | --- | --- | --- | --- | --- | --- | --- | --- | --- |
|  |  |  |  |  | R Square Change | F Change | df1 | df2 |  |
| 1 | .814<sup>a</sup> | .662 | .656 | 838.929287 | .662 | 118.947 | 7 | 425 | .000 |

a. Predictors: (Constant), aged\_65\_older, diabetes prevalence, Mean BMI Body Mean Index(kg/m), population density, Annual International Arrivals in million, Average Temperature in March(C), median age

### ANOVA table

The ANOVA table reports a significant *F* statistic (*F*=118.947, p<.005), which tests the null hypothesis that the predictors show no relationship to the criterion. Since the *F* statistic is significant (p<.05) the null hypothesis is rejected which means that the predictors (Risk factors) show a significant relationship to the criterion (Vulnerability to COVID-19) or in other words the multiple regression is overall significant.

Last, the table provides us with the data we need to compute R^2^. If we compute SS-regression divided by SS-Total, we get the R^2^.

**R**^**2**^ **= SS-regression/SS-Total = 586005052**.**988/885121050**.**800= .662**

**ANOVA^a^**

| Model |  | Sum of Squares | df | Mean Square | F | Sig. |
| --- | --- | --- | --- | --- | --- | --- |
| 1 | Regression | 586005052.988 | 7 | 83715007.570 | 118.947 | .000<sup>b</sup> |
|  | Residual | 299115997.812 | 425 | 703802.348 |  |  |
|  | Total | 885121050.800 | 432 |  |  |  |

a. Dependent Variable: total\_cases\_per\_million
b. Predictors: (Constant), aged\_65\_older, diabetes prevalence, Mean BMI Body Mass Index(kg/m), population density, Annual International Arrivals in million, Average Temperature in March(C), median age

### Coefficients table

Even though the overall model fit looks positive (see above), the coefficients table shows that there is a non-significant coefficient for some redictors indicating that some variables don’t significantly contribute to the model.

**Coefficients**^**a**^

| Model |  | Unstandardized Coefficients |  | Standardized Coefficients | t | Sig. | Collinearity Statistics |  |
| --- | --- | --- | --- | --- | --- | --- | --- | --- |
|  |  | B | Std. Error | Beta |  |  | Tolerance | VIF |
| 1 | (Constant) | 5001.555 | 20835.612 |  | .240 | .810 |  |  |
|  | median age | 722.846 | 312.592 | 3.968 | 2.312 | .021 | .000 | 3702.946 |
|  | population density | 2.695 | 7.139 | .088 | .377 | .706 | .015 | 67.974 |
|  | Annual International Arrivals in million | .021 | .011 | .475 | 1.829 | .068 | .012 | 84.966 |
|  | Mean BMI Body Mass Index(kg/m) | -943.494 | 1020.313 | -.512 | -.925 | .356 | .003 | 384.927 |
|  | diabetes prevalence | -96.082 | 308.948 | -.120 | -.311 | .756 | .005 | 188.003 |
|  | Average Temperature in (C) | 236.612 | 109.753 | .766 | 2.156 | .032 | .006 | 158.808 |
|  | aged_65_older | -699.974 | 293.445 | -3.317 | -2.385 | .017 | .000 | 2432.349 |

a. Dependent Variable: total\_cases\_per\_million

If we look at the standardized coefficients, **median age** actually contributes more to the model because it has a larger absolute standardized coefficient (**β = 3**.**968**). Aged 65 and plus contribute larger to the model with an absolute standardized coefficient (**β = 3**.**317**)

The regression equation of the model is:

**Vulnerability to COVID-19 =** 5001.555 + 722.846 **× median age +** 2.695 × **population density** + .021 × **number of International arrivals per year** −943.494 × **Body Mass Index** - 96.082 × **diabetes prevalence +** 236.612 × **average temperature in (C)** - 699.974 × **aged plus 65**

### Correlations able

**Correlations**

|  |  | total_cases_<br>per_million | median<br>age | population<br>density | International<br>Arrivals in M | Mean<br>BMI | Diabetes<br>preva-<br>lence | Tempe-<br>-rature<br>March<br>(C) | age><br>65 |
| --- | --- | --- | --- | --- | --- | --- | --- | --- | --- |
| Pearson<br>Correlation | total_cases_<br>per_million | 1.000 | .563 | .478 | .609 | -.467 | .146 | -.468 | .533 |
|  | median_age | .563 | 1.000 | .663 | .510 | -.448 | .038 | -.839 | .991 |
|  | population_<br>density | .478 | .663 | 1.000 | .498 | -.633 | -.175 | -.690 | .692 |
|  | Annual<br>International<br>Arrivals in<br>million | .609 | .510 | .498 | 1.000 | -.645 | -.310 | -.679 | .537 |
|  | Mean BMI<br>Body Mass<br>Index<br>(kg/m) | -.467 | -.448 | -.633 | -.645 | 1.000 | -.236 | .655 | -.499 |
|  | diabetes_pre<br>valence | .146 | .038 | -.175 | -.310 | -.236 | 1.000 | .163 | -.010 |
|  | Average<br>Temperature<br>in March(C) | -.468 | -.839 | -.690 | -.679 | .655 | .163 | 1.000 | -.855 |
|  | aged_65_old<br>er | .533 | .991 | .692 | .537 | -.499 | -.010 | -.855 | 1.000 |
| Sig.<br>(1-tailed) | total_cases_<br>per_million | . | .000 | .000 | .000 | .000 | .001 | .000 | .000 |
|  | median_age | .000 | . | .000 | .000 | .000 | .215 | .000 | .000 |
|  | population_<br>density | .000 | .000 | . | .000 | .000 | .000 | .000 | .000 |
|  | Annual<br>International<br>Arrivals in<br>million | .000 | .000 | .000 | . | .000 | .000 | .000 | .000 |
|  | Mean BMI<br>Body Mean<br>Index(kg/m) | .000 | .000 | .000 | .000 | . | .000 | .000 | .000 |
|  | diabetes_pre<br>valence | .001 | .215 | .000 | .000 | .000 | . | .000 | .420 |
|  | Average<br>Temperature<br>in March(C) | .000 | .000 | .000 | .000 | .000 | .000 | . | .000 |
|  | aged_65_old<br>er | .000 | .000 | .000 | .000 | .000 | .420 | .000 | . |

From the table of correlations above, we can see that all the risk factors considered in this study are statistically correlating with the dependent factor (with different degrees): total infected COVID-19 in-hospital cases for all the seven countries with sig ‹.005. As expected, median age and percentage of aged more than 65 years old are strongly correlated with a r more than 0.5. Temperature is negatively strongly correlated with the vulnerability to COVID-19 with r= −0.468. Annual international arrival and population density are also significantly positively correlated with the vulnerability to COVID-19. However, diabetes prevalence, and as expected, is not significantly associated with Vulnerability to COVID-19 with r= 0.146.

## CONCLUSION

In this article we highlight the causality between seven risk factors (social, demographic, climatic, public health) and vulnerability to COVID-19, the exploration shows that several factors do not act alone and that their correlation with the vulnerability is very low, we cite in particular the weight (body mass index, BMI) and the prevalence of diabetes. International arrivals seem to be a facilitator of the spread of the virus. Thus, based on international arrivals in millions for each country, also and on the basis of the data, we can support a direct correlation between the total number of passengers and the total number of infected cases. We find a strong positive correlation between the percentage of people over the age of 65 and the total number of infected cases for almost all countries except Greece, where more than 20% of its population is over the age of 65. COVID-19 appears to be a rhomboid age pyramid disease.

The population density effect seems to increase the number of infected individuals and increase the risk of vulnerability and death within clusters without being the main cause.

Population density can vary by a coefficient ranging from 1 to 4 in the 7 countries on our panel. However, when it is low, the number of infections increases much less quickly and this would be one of the indicators explaining the resilience of some countries to coronavirus.

The ratio of people over 65 is three times higher in the five European countries than in South Africa and In Morocco, the population density is 2 to 4 times higher in these same five countries than in the two African countries. That is said, when these two indicators are weak and do not increase the number of infected individuals, they would be essential indicators including the improvement of the health resilience of Morocco and South Africa in the face of the Coronavirus. Note that the number of deaths in Morocco and South Africa is only 0.7% that of the three most affected European countries Spain, Italy, France. We are not talking about systemic resilience related, for example, to the emergency health, security and social measures taken by these two countries in terms of lockdown, public policy choices and socio-economic measures.

To our knowledge, this comparative study is considered to be the richest in terms of number of countries and individuals including Africa and Europe. For most countries the bed capacity, the number of ventilators and the basic reproductive rate (R0) of COVID-19 were the three key instruments for steering this health crisis. Those who have managed to control them efficiently have been able to show strong health and social resilience. We noted that in the countries in our sample the lockdown of the entire population logically contained the spread of the virus and reduced vulnerability.

Ultimately, the resilience of health systems to pandemics is not only dependent on variables affecting health vulnerability, but other exogenous and qualitative factors that can either make it strong or more fragile. That’s way, the implementation of emergency public policies such as border closures, food autonomy, decentralization of health resources, reduction of political debates and bureaucratic constraints have made managing the pandemic more practical and easier.

## Data Availability

we declare the availability of all data refered to in the manuscript.
https://www.weatherbase.com/weather
https://www.worldometers.info/
https://ourworldindata.org/coronavirus-source-data
https://www.who.int/emergencies/diseases/novel-coronavirus-2019

https://www.weatherbase.com/weather

https://www.worldometers.info/

https://ourworldindata.org/coronavirus-source-data

https://www.who.int/emergencies/diseases/novel-coronavirus-2019

## Data Sources

https://www.weatherbase.com/weather

https://www.worldometers.info/

https://ourworldindata.org/coronavirus-source-data

https://www.who.int/emergencies/diseases/novel-coronavirus-2019

